# Canadian newspapers support mifepristone medical abortion to improve fulfillment of the right to health (2015-2019)

**DOI:** 10.1101/2022.07.11.22277487

**Authors:** Tamil Kendall, Pallavi Sriram, Amrit Parmar, Wendy V. Norman

## Abstract

In 2015, mifepristone, the international gold standard for medical abortion, was approved for use in Canada. Our content analysis of Canadian newspaper coverage describes arguments in favor or against medical abortion and the evolving regulatory framework for mifepristone from six months before approval until the last province included the medication as a publicly insured health benefit (2015-2019).

Our study found an exceptionally high level of support for the approval, introduction and removal of regulatory barriers to mifepristone for medical abortion. Of 402 articles, 67% were pro-medical abortion, 25% presented balanced or neutral coverage and only 8% presented solely anti-medical abortion viewpoints. Of the 761 stakeholders cited, more than 90% made positive or neutral statements about medical abortion. Most articles discussed medical abortion as a health issue and contained arguments about how liberalization of the regulatory framework and public payment for mifepristone would improve abortion availability (68%), accessibility (87%), acceptability (34%) and quality (19%). Mifepristone’s introduction in Canada was presented in newspapers as a way to increase women’s health, specifically in rural areas where disparities in abortion access exist.

Rather than formal balance, which presents contrasting arguments as equally valid even when the scientific evidence for one vastly outweighs the other, our study identified evidentiary balance where coverage aligned with the majority of evidence and expert opinion. Our results differ from analyses in other high-income countries (United Kingdom, United States) that have found that media frame abortion as a stigmatized and controversial issue or mention abortion predominantly with respect to electoral politics rather than as a health issue. The Canadian print media presented overwhelmingly favorable arguments towards the expansion of mifepristone medical abortion and served to destigmatize abortion by framing the introduction and universal coverage of medical abortion as fulfilling core components of the right to health.

## Introduction

In 2015 Health Canada, the federal regulator, approved mifepristone for medical abortion in Canada. Mifepristone, also known by the name RU-486, is the gold standard medication for medical abortion^1^ according to the World Health Organization (WHO) ^2,3^. It was unavailable in Canada until January 2017, when it reached the market as Mifegymiso ®^4^, a combination pack of mifepristone and misoprostol.^5^ The introduction of mifepristone is arguably the biggest national policy change to affect the delivery of abortion services in Canada since 1988 when the Supreme Court of Canada found that the criminalization of induced abortion was unconstitutional and abortion was struck from the criminal code, subsequently to be treated like any other medical procedure.^6,7^

Canada is a provincially decentralized federation where the federal government is responsible for regulation of medical devices and pharmaceuticals, but provinces are responsible for the regulation, coverage and delivery of public healthcare services in alignment with minimum universal standards. The federal statute, the Canada Health Act, ^8^ sets out the inter-relationships and responsibilities. For decades, abortion has been a legal^9^ and common^10^ healthcare procedure in Canada, with nearly one in three women having an abortion during their lifetime.^11^ Nevertheless, rural-urban disparities in abortion access have been documented in Canada.^12,13^ In 2016, the United Nations Committee on the Elimination of Discrimination Against Women expressed concern about disparities in access to abortion and called upon Canada to “ensure access to legal abortion services in all provinces and territories”.^14^

As a highly effective form of medical abortion that requires less provider specialty training and infrastructure,^3,15^ mifepristone has the potential to address the rural-urban disparity in abortion access in Canada.^16^ However, initial approval in July 2015 was accompanied by significant regulatory barriers to access such as physician-only dispensing and direct observed dosing of the medication.^17^ Experience in other countries, including France and the United States of America (USA), indicates that over and above the approval of mifepristone for use in medical abortion, the regulatory framework for its distribution influences the rapidity and extension of use after introduction.^18^

The period between the federal regulatory approval and last provincial approval for universally subsidy of mifepristone represents a unique historical moment. We chose to examine this moment to understand how the issue of mifepristone specifically and medical abortion in general was represented by the news media. This period encompasses six months leading up to the approval of mifepristone for use in Canada in July 2015, the subsequent modifications of the regulatory framework (including the removal of most barriers to prescribing and dispensing by November 2017)^19,20, 21^ as well as the decisions by provincial health plans to include mifepristone as a covered health benefit with the last province doing so in 2019.

The press is a medium for politicians and interest groups to express their views. This contributes to setting the policy agenda by influencing public attention and understanding of issues. These expressions in the press also influence perceptions of priority, potential policy responses and, at times, actions of policymakers.^22-24^ We were interested in how the introduction and regulation of mifepristone for medical abortion was reported by Canadian print media, with a specific focus on arguments made for and against regulations and coverage by provincial health plans as they related to the four components of the right to health: availability, accessibility, acceptability and quality.^25-27^

## Methods

We performed a content analysis of all Canadian newspaper articles published from January 1, 2015 to November 30, 2019 including the terms “medical abortion” or “abortion pill”. We identified articles by searching the Canadian Newsstream^28^ database which includes the full text of more than 360 Canadian Newspapers from across the country, representing the major daily newspapers as well as smaller community newspapers. We chose the time period from January 1, 2015 to November 30, 2019 to include the time leading up to the approval of mifepristone in July 2015 until after the last Canadian province included mifepristone as an insured benefit in September 2019. Two of Canada’s three territories include mifepristone as an ensured health benefit (Yukon since October 2018 and Northwest Territories since June 2019). The third territory, Nunavut, does not include mifepristone as an ensured health benefit but more than 90% of Nunavut’s population has been insured for mifepristone since May 2017 through the federal governments’ non-insured health benefit program for eligible First Nations and Inuit people.^29,30^

### Sample selection

We read full text record in full to determine relevance. We excluded articles if they:

a. did not discuss the introduction and regulation of medical abortion in Canada;
b. were not a newspaper or newswire story (for example the record was transcribed dialogue of a TV interview); or
c. were an exact duplicate copy abstracted twice in the database (same text published with the same title by the same outlet on the same day).

We treated variations of stories or editorials that were substantially similar, for example a newswire story published by different news outlets with different headlines and minor editorial changes and print and online versions of a story that included different content published by the same news outlet (for example a shorter print and longer online version of an article), as unique records.

### Coding

Our content analysis of the newspaper articles applied an *a priori* coding scheme to identify the main themes and categorize the article’s tone (pro-medical abortion, neutral or anti-medical abortion). We tested and validated the coding framework by two coders coding a sub-sample of 10 articles and comparing their results. Emergent themes and questions that arose during coding were discussed and agreed upon by AP and TK.

If the article included quotes from individuals that expressed pro-medical abortion and anti-medical abortion views, we coded the article as neutral. If the article did not include any arguments for or against medical abortion, for example the announcement of inclusion of mifepristone coverage in a provincial drug plan without any editorial commentary from the journalist or expression of pro or anti positions, we coded the article as neutral. If the journalist, editor or letter writer and/or the individuals quoted were exclusively supportive of introduction of mifepristone in Canada and removal of regulatory barriers to medical abortion, including coverage of mifepristone in provincial drug plans, we coded the article as pro-medical abortion. If the journalist, editor or letter writer only expressed arguments against mifepristone or abortion in order to critique them, we coded the article as pro-medical abortion. If the journalist, editor, letter writer and/or individuals quoted expressed only opposition to the introduction of mifepristone for medical abortion and removal of regulatory barriers we coded the article as anti-medical abortion.

We categorized the themes using the core components identified in the WHO’s AAAQ (Availability, Accessibility, Acceptability, and Quality) right-to health framework,^25^ identifying *availability* (abortion availability and discussion of regulations that would limit uptake and availability specifically physician and ultrasound availability, additional training for healthcare providers and physician dispensing), *accessibility* (financial and geographic barriers in the context of mifepristone introduction e.g. provincial insurance coverage, travel time to point of care), *acceptability* (privacy, cultural acceptability, restrictive regulations that could reduce acceptability, such as physician observed dosing, or increased administrative burden such as patient written consent) and *quality* (medically approved, scientifically valid and of good technical quality). With respect to quality, we analyzed whether the article provided information about the administration and function of mifepristone as well as arguments made about safety and health risks. We identified whether information from reputable medical sources, such as the WHO or the Society for Obstetricians and Gynecologists of Canada, was provided. We also specifically coded for and analyzed arguments made against abortion (anti-abortion discourse) and discussion of abortion as a human or women’s rights issue.

We further analyzed which professional categories of stakeholders are cited in the articles: academics/researchers (who were labelled as such by the journalist or affiliated with a university), health professionals (coded by individual profession e.g. physician, midwife, nurse, pharmacist and then grouped together), elected or staff representatives of government, civil society organizations (such as Action Canada for Sexual and Health and Rights or Campaign for Life), religious leaders, and members of the public. During coding, we added the category “abortion clinic staff” to identify individuals who are not healthcare providers and who were supporting provision of abortion care in either a public or not-for-profit setting. The professional category was determined by the first mentioned professional designation or occupational sector, or affiliation given for a stakeholder who was quoted in the article. For example, a stakeholder described as a professor or researcher or affiliated with a university would be coded as an academic. An individual quoted in more than one article is coded for each article they were cited in. For each article, the professional category of each stakeholder quoted, and tone of the statement (pro-medical abortion, anti-medical abortion and neutral) was coded.

## RESULTS

Of the 600 records for news articles published between January 2015 and November 2019, we included 402 in the analytic sample. Of all 600 records, we excluded 198 because they were not relevant (did not address the introduction and regulation of medical abortion in Canada), were not a newspaper or newswire story, or were an exact duplicate. Of the 402 articles included in the sample, 113 were variations of the same story with minor variations in the content.

### Article tone and stakeholders quoted

In Canadian print media coverage of the introduction and regulation of mifepristone between 2015 and 2019, we found the majority of articles were pro-medical abortion (269 articles, 66. 9%), followed by neutral coverage (102 articles, 25.4%), see Figure 1. Only 31 articles (7.7%) expressed solely negative attitudes towards the introduction of, and increasing access to, medical abortion in Canada (labelled anti-medical abortion). The vast majority of the anti-medical abortion articles were letters to the editor (30 of 31); one newspaper story on political candidates running as independents included a negative statement about mifepristone by a candidate running on an anti-choice platform.^31^ Interested in the concentration of anti-medical abortion views in opinion pieces, we analyzed the distribution of anti, neutral and pro-medical abortion views expressed in opinion pieces (letters to the editor, op eds and editorials). Of the 67 opinion pieces, 33 (49.2%) were pro-medical abortion (including 17 out of 18 editorials published by editorial boards with the 18^th^ editorial taking a neutral stance); 30 (44.8%) were anti-medical abortion and these were all letters to the editor from the public; and 4 opinion pieces (6.0%) presented arguments both in favor and against expansion of medical abortion in Canada through the introduction and regulation of mifepristone, and were coded as neutral.

**Figure 1:**
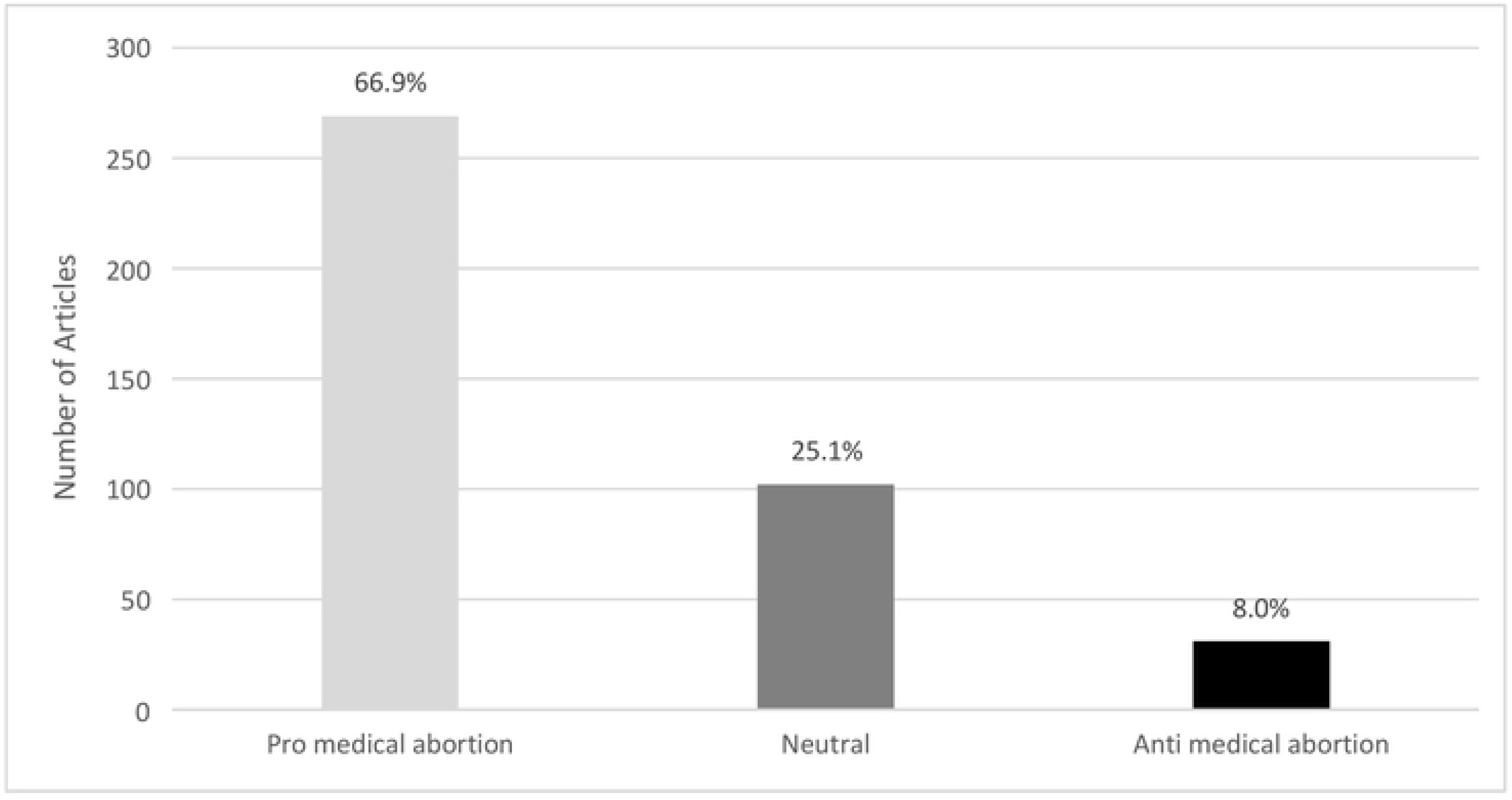
Canadian newspaper articles by tone of reporting, Jan 2015 to Nov 2019.

#### Frequency of anti, neutral and pro-medical abortion statements and stakeholder professional categories

In total, there were 761 quotes from stakeholders in our sample of 402 articles. The frequency of anti-medical abortion statements made in the sample was 9.3% (71 of 761), neutral 16.3% (124 of 761) and pro-medical abortion 74.4% (566 of 761).

The most common professional categories of quoted speakers were government (218, 30.3%), civil society organizations (189, 23%) and healthcare professionals (129, 17.9%), see Figure 2. Within the professional categories, the voices of healthcare professionals (93.7% of physicians, 92.3% of nurses) and academics (94.5%) were predominantly in favour of the introduction of, and increased access to, mifepristone in Canada. Pro-medical abortion views also dominated those solicited from speakers affiliated with abortion clinics (90.3% pro and 9.7% neutral) and organized civil society (76.2% pro). The groups which held the largest proportion of perspectives against medical abortion were the public (22.9%), organized civil societies (19.8%) and the government (11.0%). However, the majority of voices within each of these groups were still pro medical abortion (public [62.9% pro], organized civil society [77.0% pro] and government [60.6% pro]). There were 2 religious leaders quoted in the pool of 761 quotations, both of whom made anti medical abortion statements.

**Figure 2:**
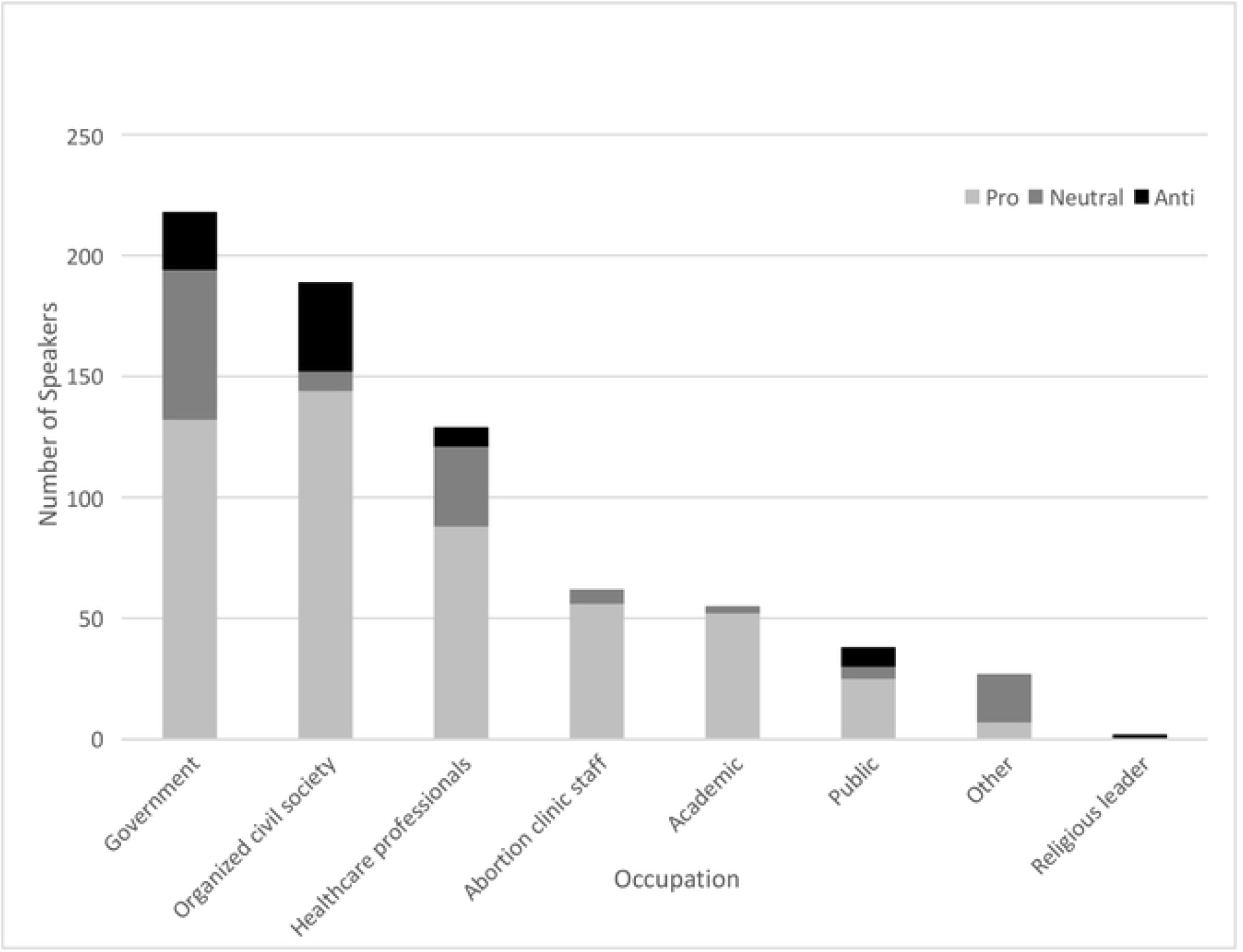
Speakers in Canadian newspaper mifepristone articles by occupation and tone, Jan 2015 to Nov 2019.

### Thematic Analysis using the AAAQ Framework

Each news article was coded to analyze the main themes presented. Of the 402 articles included in this study, almost half (190, 47.15%) identified one or more regulatory, financial and/or health system barriers to medical abortion. We apply the WHO’s AAAQ right to health framework^27^ to categorize the types of barriers or facilitators of medical abortion identified in Canadian print media and thematically analyze and illustrate the types of arguments made by different stakeholders.

#### Availability

In Canada, print media coverage of the approval and introduction of mifepristone for medical abortion put a clear emphasis on the issue of availability. First, the drug regulatory process in Canada which approved mifepristone in 2015 was identified as an important step to improve the availability of medical abortion in Canada in 25 articles (6.2%). For example, the CP Wire service in 2015 stated:

> “*To be clear, medical abortion has been available in Canada for decades. However, Canadian women have had to rely on less effective, less safe medications for this purpose. This is a step in the right direction towards ensuring Canadian women have access to legal first trimester abortion care that is as safe and accessible as it is in other parts of the world*.” (30/07/2015)

More than half of the articles reviewed (68.2%, n=274) discussed how increased access to mifepristone would support increased abortion availability. A third of these (n=94, 34.3%) specifically mentioned mifepristone’s potential to increase availability of abortion in rural areas. An illustrative quote from the CP Wire Service was from Brian Gallant, then-premier of New Brunswick, the first province in Canada to introduce universal health coverage for the nearly $400 cost of Mifegymiso, saying:

*“‘We believe access to essential medical services shouldn’t depend on a person’s postal code or income bracket,’ […] Mifegymiso […] will help improve women’s access to reproductive health care, especially in rural regions far from hospitals that perform surgical abortions*.*’”*. (24/07/2017).

The specific regulations that could hamper availability of mifepristone were identified in 92 articles. The most common barriers to availability identified were requiring physicians and pharmacists to undergo special training to prescribe or dispense mifepristone (n=28, 30.4%) which was removed in May 2017, requiring physicians to be registered and certified to provide mifepristone (n=11, 11.9%) and to directly dispense mifepristone to patients (n=38, 41.3%), restrictions which were removed in November 2017, and requiring ultrasound prior to medical abortion (n=34, 36.9%) which was removed in April 2019, see Figure 3.

**Figure 3:**
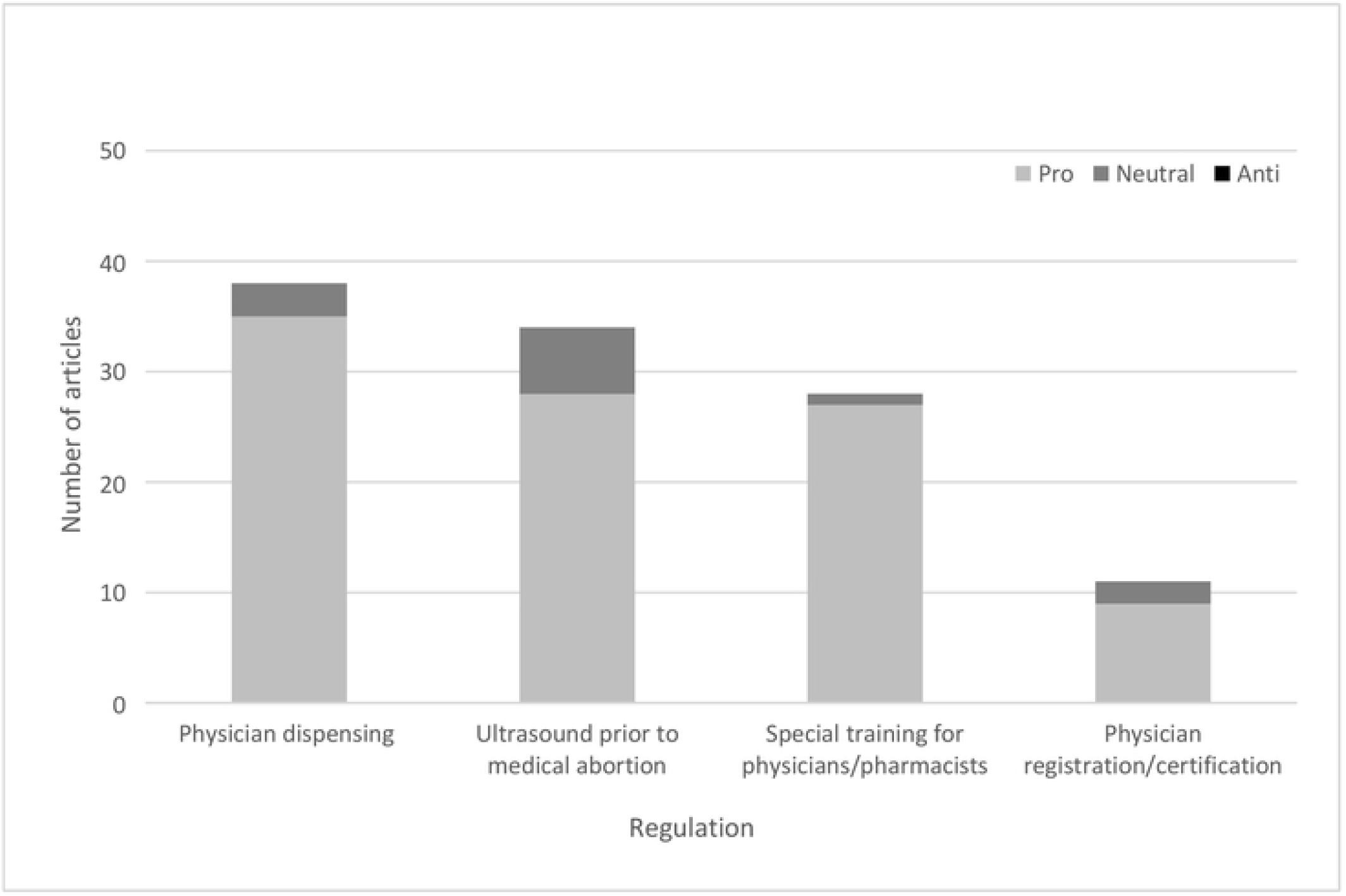
Restrictive regulations identified in Canadian newspaper articles discussing availability of medical abortion, Jan 2015 to Nov 2019. Healthcare professionals where speakers identified as physicians, nurses, pharmacists or emergency service providers

When mifepristone was first approved, regulations required physician prescribers to take an online certification course and for them to be registered with the manufacturer in order to prescribe the medication. This was identified as a time-consuming hurdle which would limit the number of health professionals who would be able to prescribe mifepristone. The Toronto Star noted that:

> “*Doctors and pharmacists, the very people unlikely to have spare work hours, must take a six-hour online course to prescribe or dispense it. Those who have taken the course are acting out of professionalism and human decency. But why should abortion still live in the realm of charity? It is legal. It is a basic human right*.” (06/03/2017).

Physician dispensing, meaning that doctors had to stock and sell mifepristone themselves rather than write a prescription that could be filled at pharmacy, was described as:

> “… *so complicated that [doctors] don’t bother with it at all. It’s so problematic… it requires a lot of jumping through hoops, something that isn’t required for any other legal prescription drug*” (Toronto Star, 07/03/2017).

Additionally, physician dispensing was discussed as a barrier to medical abortion moving into the realm of primary care because, as opposing to pharmacist dispensing, it would:

> “…*discourage family doctors who do not specialize in women’s reproductive services from offering the pills…”* (Globe and Mail, 21/01/2017) because *“It’s quite an investment for a physician to be a dispensing practitioner…” (Globe and Mail, 17/10/2016)*.

This limitation would be felt: *“*…*particularly among women in rural and remote communities whose family doctors do not have the capacity to run a parallel drug-dispensing operation*.*”* (Globe and Mail, 12/07/2016).

Obligatory ultrasound prior to medical abortion was identified as disproportionately affecting rural women. The Globe and Mail quoted an academic and health professional from Ontario saying saying:

> *“*… *women in areas where ultrasounds are not readily available, such as Nova Scotia and some remote and rural parts of Canada, may be facing unnecessary delays. These regulations are being used in a scapegoating manner to justify practice restrictions*…”

and the article identified other evidence-based methods of pregnancy dating such as pelvic exams (27/09/2018). Removing the ultrasound requirement had the potential to: “…*encourage more health providers to start prescribing Mifegymiso. It’s going to improve the process overall*.” (Globe and Mail, 17/04/2019).

#### Accessibility

The accessibility domain requires health services to be accessible to everyone without discrimination, which includes lack of access based on financial status. At least one theme related to mifepristone accessibility was discussed in the majority of articles analyzed (n=351, 87.3%). The most common barriers to accessibility identified were lack of provincial drug plan coverage for mifepristone (n=189, 47.0%) and the overlapping issue of financial access as a barrier (n=76, 18.9%). As noted earlier, drug plan coverage of mifepristone was discussed as a way of increasing availability of medical abortion, however the lack of coverage was identified as a barrier to access. For example, the Globe and Mail reported that:

> “*[m]ost women are not going to pay out of pocket [for a pill] when they can have a safe and effective surgical abortion at no cost to [them]*…*It diminishes the promise of opening access to women in more rural areas where surgical abortions are not readily available*.” (2016).

Almost half of the articles (n=200, 49.6%) included statements (either by the writer or by stakeholders cited in the article) arguing for drug plan coverage of mifepristone as a means of supporting accessibility of medical abortion. When mifepristone was initially approved for medical abortion, the cost of the medication ($300-450 CAD)^32^ had to be borne by the woman seeking it. In contrast, surgical abortion, an in-hospital or in-clinic procedure, was covered by public health insurance in Canada. Implementing drug plan coverage of mifepristone would increase accessibility of abortion, and this often overlapped with increasing rural access to abortion. For example, the Globe and Mail quoted an academic health professional from British Columbia as saying:

> “*Being able to have the medication also covered by the government, this helps tremendously so that women can access care in their own communities and can judge whether medical or surgical abortion is better for them based on what their health needs are, not what their postal code is*” (03/01/2018).

The timeline in Figure 4 describes print media coverage of the introduction and regulation of mifepristone for medical abortion in Canada from the six months prior to approval by Health Canada (January 2015), through the time when the product became available on the market in July 2017, and over the period of the removal of each of the regulatory restrictions that presented barriers to availability and accessibility, including affordability, up until the last Canadian province included coverage of mifepristone in it’s provincial pharmaceutical formulary in September 2019.

**Figure 4:**
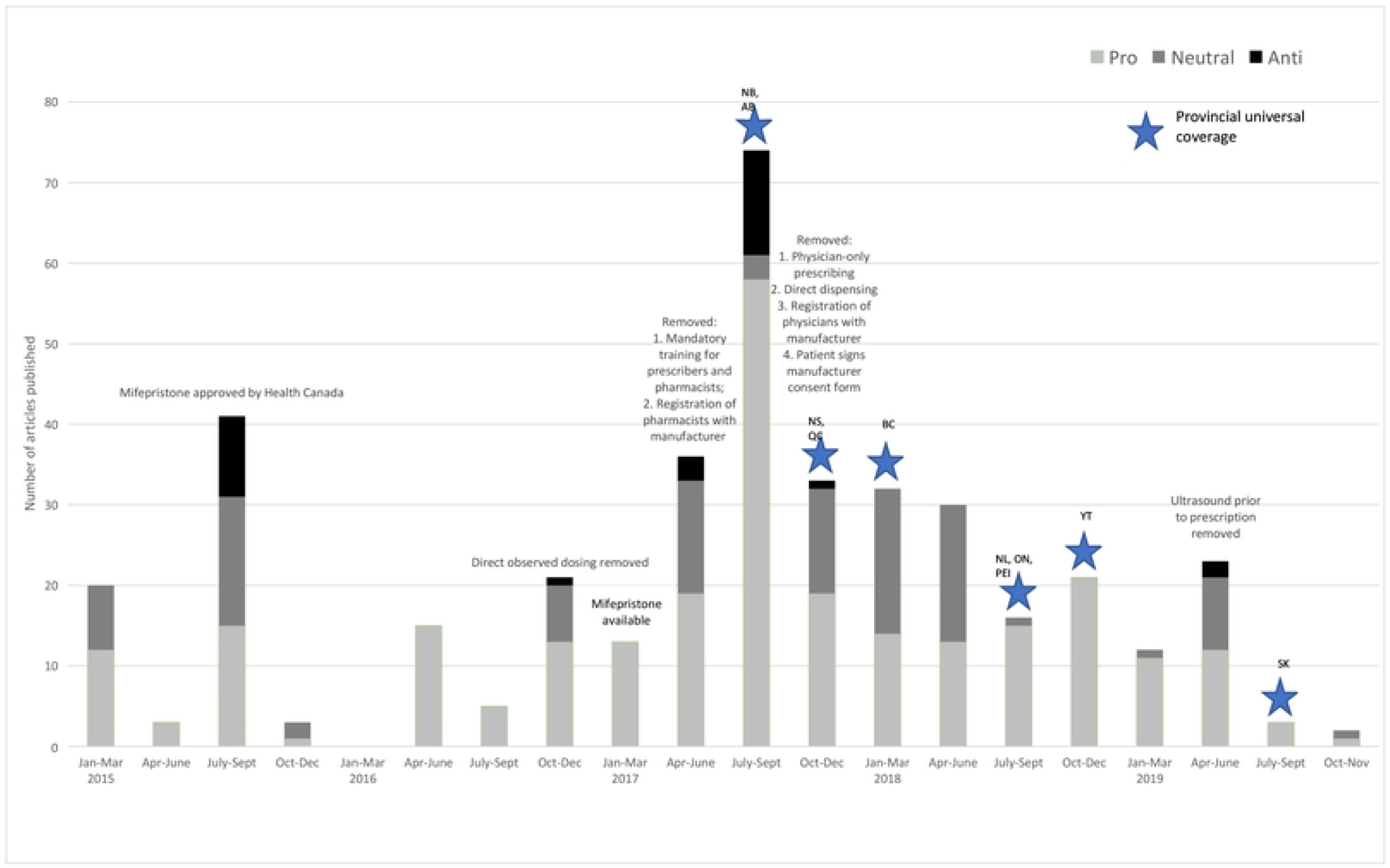
Timeline of Canadian newspaper articles published during mifepristone introduction and regulation in Canada, Jan 2015 to Nov 2019.

Figure 4 illustrates that throughout this period, the tone of print articles about mifepristone and medical abortion in Canada have been predominantly pro-medical abortion or neutral. The majority of anti-medical abortion articles printed coincided with the initial Health Canada approval of mifepristone in July 2015 and subsequently when it became available on the market in January 2017. Even at these points in time, the pro-medical abortion and neutral articles make up the majority of newspaper publications. After November 2017, when all restrictive regulations except the ultrasound requirement had been removed, only one anti-medical abortion article was published, and it coincided with the removal of the requirement for ultrasound to be performed prior to medical abortion in April 2019.

#### Acceptability

Acceptability states that health services must be respectful of medical ethics, including privacy and confidentiality, and be culturally sensitive. We found 34.1% (n=137) of articles included at least one theme on how mifepristone supported acceptability of medical abortion and how specific regulatory restrictions challenged its acceptability. Equity, cultural sensitivity and privacy were highlighted as benefits of medical abortion in 14.4% of articles (n=58). The Chief Executive Officer of the Society of Obstetricians and Gynaecologists of Canada was quoted as saying:

> “… *allowing Canadian women access to mifepristone would have no impact on the number of women choosing abortion but would help make an “intensely personal issue” a private health matter between a woman and her doctor*.” (Vancouver Sun, 13/01/2015).

The ability to offer a medical abortion at a time and place determined by the pregnant person, and thus to make the decision truly private, was discussed in 10.2% of articles (n=41). Regulations such as physician observed dosing and requiring patient written consent prior to medical abortion were identified as barriers to the acceptability of mifepristone for medical abortion in 20 articles. Opposition to physician observed dosing, the requirement for a doctor to witness women taking the mifepristone in their office, was objected to in 17 articles. A quote from an academic health professional from a western Canadian university exemplifies these arguments that physician observed dosing is: “…*medically unnecessary and demeaning to Canadian women*” (The Province, 20/04/2016).

#### Quality

Quality as a domain of the right to health, states that health services must be scientifically and medically approved. Mifepristone is considered the gold standard in medical abortion by the WHO and is on its list of essential medications.^33^ We found 19.2% of articles (n=77) included scientific information on how mifepristone works and 11.2% (n=45) of articles provided safety information from the WHO or other reputable scientific bodies.

Within the group of speakers quoted in the articles, we looked at whether journalists were using speakers with the professional background to reinforce that mifepristone is a scientifically and medically approved method of medical abortion. We specifically looked at the representation of two groups with expertise on the topic of medical abortion in Canada:

(1) The 16 physician members of the Society of Obstetricians and Gynecologists of Canada (SOGC) who authored the Medical Abortion Clinical Practice Guideline released in 2016 as subsets of all physicians quoted and (2) the 28 Collaborators of the Contraception and Abortion Research Team (CART-GRAC) working on a national study on Mifepristone Implementation in Canada (CART-Mife Study), as subsets of all of the stakeholders quoted. The SOGC is the national specialty organization for the practice of obstetrics and gynecology in Canada and is responsible for producing national clinical practice guidelines and maintaining continuing education in women’s sexual and reproductive health. CART-GRAC is a Canadian multi-disciplinary research team based at the University of British Columbia (UBC) focused on conducting national, cross-sectoral research to improve family planning care available to Canadians.

Of the 761 quotations across all articles, 8.1% (62 quotations) were from the authors of the SOGC guideline on Medical Abortion and 18.1% (138 quotations) were from people affiliated with the CART-Mife Study. Of the 313 articles which included a quotation, 18.2% (57 articles) quoted at least one speaker who authored the SOGC guideline on Medical Abortion and 30% (94 articles) quoted at least one speaker affiliated with the CART-Mife Study (Figure 5). Of the 146 quotations identified as made by physicians, 42% were authors of the SOGC guideline on Medical Abortion (Figure 6).

**Figure 5:**
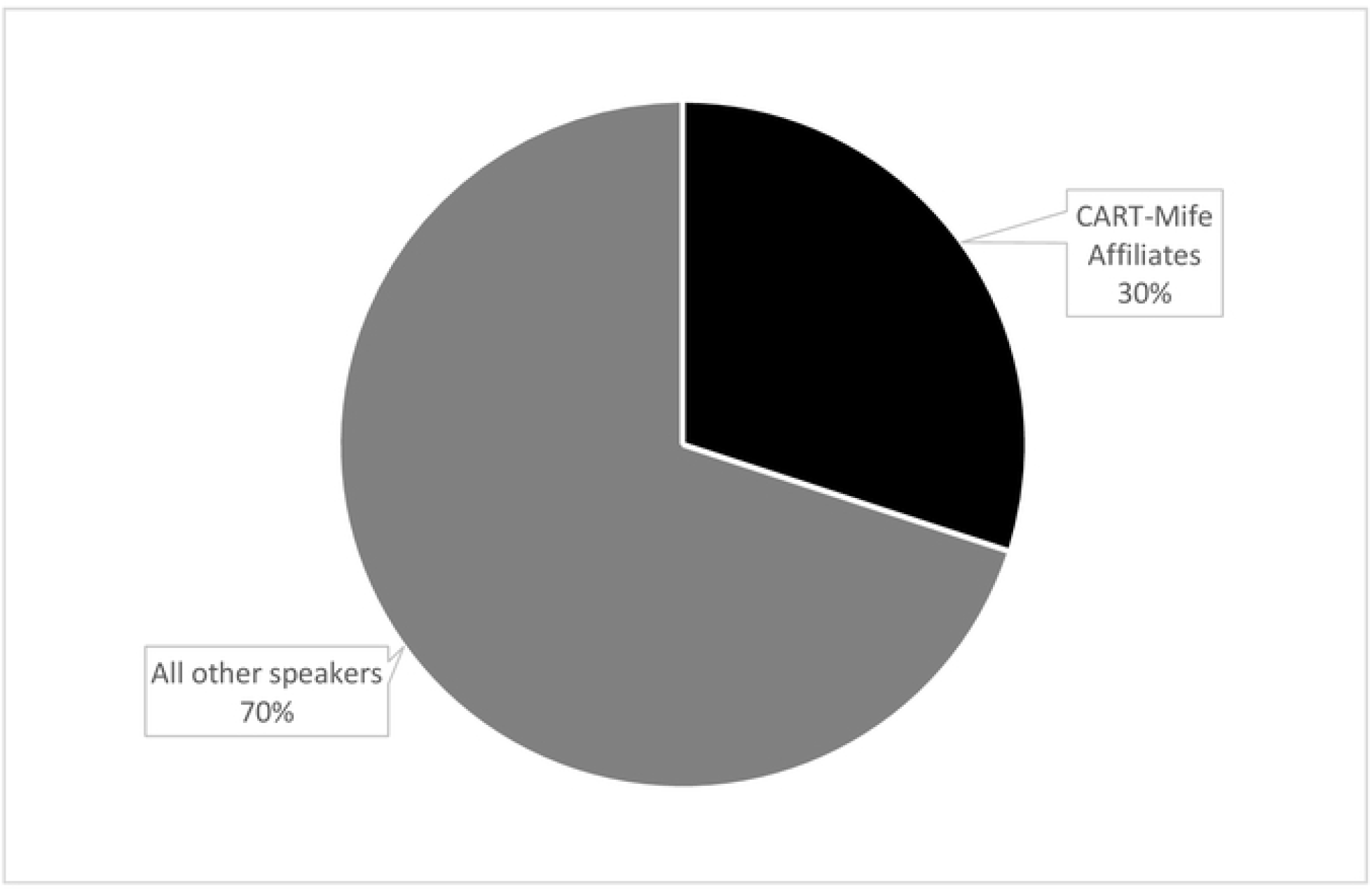
Speakers quoted in Canadian newspaper articles about mifepristone in Canada by affiliation with the CART-Mife research team, Jan 2015 to Nov 2019.

**Figure 6:**
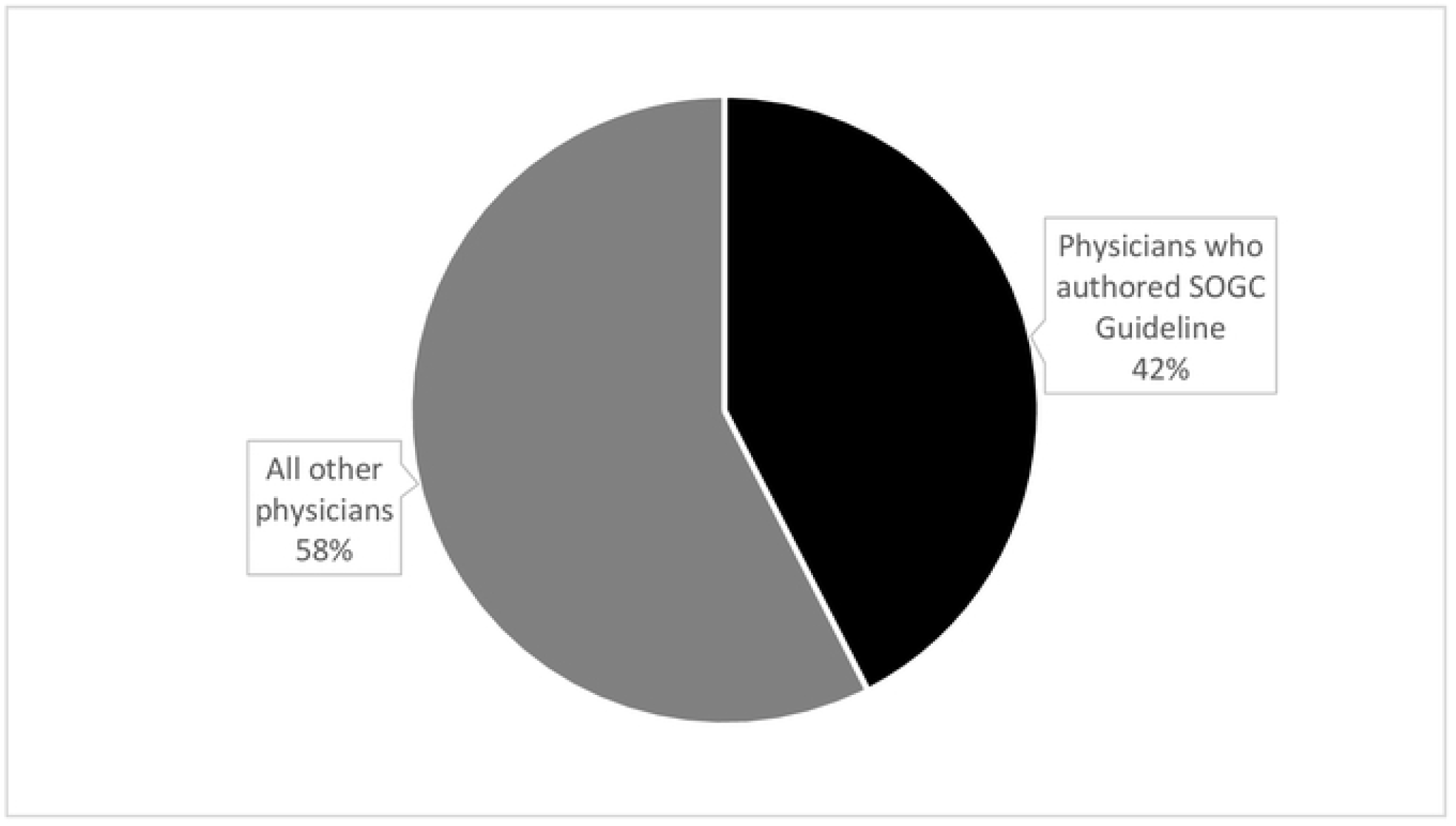
Speakers quoted in Canadian newspaper articles about mifepristone in Canada by affiliation with the SOGC abortion guideline committee, Jan 2015 to Nov 2019.

### Discussions of “the right to life”, women’s rights and personal views influencing policy in press coverage of the introduction and regulation of mifepristone for medical abortion

#### Anti-abortion discourse

We coded all of the articles for the presence of any anti-abortion discourse, which was identified in just under a fifth of articles (n= 78, 19.4%). The majority of articles that included anti medical abortion themes were news stories (n=42, 53.8%) and letters to the editor (n=30, 38.5%). The others were wire stories (n=5, 6.4%) and 1 editorial (1.3%). More than half of the articles that expressed any anti-abortion views also included statements from supporters of increased abortion access or only quoted anti-medical abortion themes to critique them (all wire stories, the editorial and 41 of 42 news stories were neutral in tone). Only letters to the editor from the public made solely anti-medical abortion arguments and were categorized as anti-medical abortion in tone.

The most common arguments made against medical abortion included the assertion that abortion is a form of homicide (mentioned in 38 articles), morally wrong in that it disrespects the sanctity of life or is an irresponsible choice (mentioned in 24 articles) or is harmful to women’s health (mentioned in 26 articles). Less common themes included the need to provide women with other options, including adoption in case of unintended pregnancy (mentioned in 2 articles), that medical abortion will increase the overall number of abortions (mentioned in 1 article) and opposition to public funding for medical abortion (mentioned in 12 articles).

Of the articles that included statements about the danger of medical abortion for women’s health, 23.07% (n=5) also included information on the safety of medical abortion from the WHO or other reputable national or professional scientific body and 26.9% (n=7) included scientific information about how mifepristone works. Analyzing these 7 articles as a sub-set provides insight into the overall positioning of anti-abortion discourse with Canadian print media coverage during the introduction and regulation of mifepristone in Canada. In all but one of the articles, mifepristone is described as an “essential medicine” and reputable scientific information about how mifepristone works is provided. Mifepristone is described as inducing *“an abortion similar to a natural miscarriage within one to two days after a woman takes the pills”* in 5 of 7 articles. In 6 of 7 articles, the ratio is two or three pro-medical abortion speakers for each anti-medical abortion speaker.^34-39^ Furthermore, the pro-medical abortion speakers are highly credible because of their professions and organizational affiliations. For the most part these speakers, who attest to the safety of medical abortion and mifepristone’s potential to improve access and reproductive health outcomes, are either physicians working with prestigious hospitals and national medical organizations, or representatives of government. The sole dissenting anti-medical abortion stakeholder quoted is from a civil society organization and is not a physician.

The seventh article in the sub-sample includes information on a provincial government’s announcement of inclusion of mifepristone in the provincial formulary, as well as information about how the drug works. It features one quote each from anti and pro-medical abortion civil society organizations.^40^

#### Abortion as a woman’s right

A minority of articles (15.4%, n=62) discussed abortion as a women’s right and how mifepristone supports women to exercise this right. Government officials made statements on abortion as a woman’s right, including then and current Prime Minister Justin Trudeau who was quoted as saying “… *Canada would remain unequivocal in its defence of a woman’s right to end a pregnancy*.” (The News, 16/05/2019). Just as those expressing anti-abortion viewpoints invoked women’s health in their arguments against abortion, the framing of abortion as a woman’s right (the right to choose) was also frequently linked to the right to health and to access to healthcare among those who spoke in favor of women’s right to make their own decisions about abortion. A quote that exemplifies the parallels drawn between women’s right to choose and the right to health came from Stephen McNeil, then and current premier of Nova Scotia, who is identified in the article as a Roman Catholic. He framed his perspective when addressing the issue as that of “a parent of a daughter” before going on to state his support for the removal of restrictive regulations on medical abortion:

> *“She [my daughter] has the right to make a decision for herself and my job is to love my daughter if she so chooses to make a decision on her health […] it’s about doing what’s right and we believe women deserve access to this health care and we will do so*.” (CP Wire Service, 22/09/2017).

#### Should the personal be political?

An unanticipated emergent theme in our content analysis was discussion of the place of personal views in politics and policy-making. This theme was discussed in 25 articles. Among these, 22 articles (88%) stated that personal belief should *not* be a part of public policy decisions. These 22 articles included statements from elected government officials discussing mifepristone but declining to state their personal views on abortion in general or mifepristone specifically. For example, The Daily Gleaner reported that:

> “*Justice Minister Peter MacKay declined to comment on the decision [of Health Canada to approve mifepristone] during a stop in Calgary to make an unrelated announcement. “Oh gosh. I think that would best be directed to (Health) Minister (Rona) Ambrose,” he said. “It is an issue of course for women, families, Canadians everywhere, but I think your question would best be directed to Health Canada…Personal opinions are best kept personal*.*”*“ (31/07/2015).

The executive director of Action Canada for Sexual Health and Rights was quoted as stating: “*Health ministers are responsible for ensuring our reproductive health, and no personal or political beliefs should interfere with people claiming those rights*.” (Winnipeg Free Press, 27/06/2018). There were 3 articles (12%) in which an elected government official or a candidate who planned to stand for election said that they would advocate for restrictions on mifepristone because of their personal beliefs against abortion.

## Discussion

Our analysis of Canadian newspaper coverage of mifepristone from January 2015 to November 2019 found an exceptionally high level of support for the approval, introduction and removal of regulatory barriers to medical abortion. Further, newspapers privileged scientific and factual perspectives from experts on the introduction and regulation of mifepristone in Canada. This alignment with the weight of scientific evidence and expert opinion is an example of evidentiary, as opposed to formal balance, in media coverage of abortion.^41^

Over two thirds of articles were pro-medical abortion, that is they only made statements or stated quoted stakeholders who supported the introduction and expansion of medical abortion, with a further quarter of articles presenting balanced or neutral coverage. Purely anti-medical abortion views were marginalized both in terms of quantity (8% of the records reviewed) and in terms of quality of reporting, as almost all were letters to the editor from the public. Only a single article among the 402 (0.3%) represented a news article presenting solely an anti-medical abortion perspective. Similarly, over ninety percent of the quoted statements from stakeholders were positive or neutral. Across all sectors of stakeholders, mifepristone statements were dominantly pro-medical abortion, whether from government, civil society or healthcare providers.

In the United Kingdom, Purcell et al analyzed articles on abortion from British and Scottish newspapers in 2010.^42^ They found that the majority of articles described abortion using negative language and that abortion was presented as being ‘controversial’. Abortion was stigmatized by being associated with ‘undesirable’ traits such as binge drinking, promiscuity and teen pregnancy. The framing of abortion as a positive or legitimate choice was marginalized. In contrast we found the anti-medical abortion discourse was marginalized in Canada, with over 90% of articles in Canadian print media being classified as pro-medical abortion or neutral.

The marginalization of anti-medical abortion views within Canadian print media, both in the volume and in the location of publication (letters rather than news stories) has important implications to discuss for journalistic balance. In the context of the United States, Sisson et al found that even among journalists publishing in feminist news outlets, 65% of journalists grappled with the concepts of neutrality and balance when reporting on abortion, and a quarter felt compelled to present “both sides of the abortion debate” as a requirement of balanced reporting.^43^ Only a third of these USA reporters took a stance more similar to our findings in Canadian newspaper coverage of mifepristone: placing priority on the merit of the arguments made and not uncritically replicating misinformation.^43^ This USA study highlights the problem false balance in reporting. When an issue already has scientific consensus, presenting ‘the other side’ gives that non-scientific or ‘anti-science’ perspective an unwarranted legitimacy and implies equal standing. This can result in uncertainty and confusion among the public about the established scientific evidence.^44^

Our study of print media coverage in Canada demonstrates evidentiary balance. We found greater weight was given to evidence-based or scientific views in discussions of the introduction and regulation of mifepristone medical abortion. The distribution of abortion reporting – over 90% reporting pro- or neutral views to the introduction of a medication which would increase access to medical abortion, reflects that the weight of scientific opinion agrees that abortion is a necessary medical procedure for women’s health and that abortion is relatively safe, for example safer than childbirth.^45^ This is further supported by the choice of authority figures such as physicians and scientific researchers as quoted stakeholders in pro and neutral articles. Our analysis showed that when physicians were quoted, 42% of the physicians were noted Canadian experts in abortion, having contributed to a national guideline on medical abortion. Around one third of articles with quotations, quoted someone affiliated with a national university-based study on the use of mifepristone. By consistently including expert voices in support of mifepristone and medical abortion, and relegating the dissenting, non-expert voices to letters/opinion pieces, the Canadian media has presented a view on abortion which is balanced in that it communicates expert consensus on the issue.

Further, we found consistency over time in the tone of coverage of medical abortion. Coverage was positive from six months before the approval of mifepristone through to announcement of public coverage by the last province. Over the study period, Canadian provincial and federal regulators and governments acted to increase the availability, accessibility and acceptability of medical abortion in Canada. They iteratively liberalized regulations and policies resulting in a federal system with the least restrictive regulatory framework for mifepristone medical abortion in the world.^6,46^ The Canadian print media communicated arguments that supported these regulatory and policy changes as opposed to arguments that could create backlash against the establishment of mifepristone medical abortion is available as an integrated part of the publicly funded, universal healthcare system.

The rural-urban disparity in abortion access in Canada has been repeatedly identified by Canadian researchers,^12,13,47-52^ with mifepristone access via primary care being proposed as a solution to improve access in rural areas.^16^ This was also the narrative presented by the Canadian media. Most articles argued for changes to the regulatory framework which would support increased availability, accessibility and acceptability of mifepristone medical abortion. When themes were analyzed from the AAAQ right to health^25-27^ perspective, articles consistently presented mifepristone as a way to support the availability (68.2%, n=274), accessibility (87.3%, n=351) and acceptability (34.1%, n=137) of medical abortion for all. A third of articles that addressed the issue of availability specifically focused on mifepristone increasing availability of rural abortion. Among the 313 articles which included speakers, researchers with the CART-Mife team were quoted 30% of the time and physicians on the SOGC’s Medical Abortion guideline committee were quoted 18.7% of the time. Citation of these authorities by the media reinforced that mifepristone is of high quality, scientifically accepted and medically approved medication for medical abortion.

Our finding that mifepristone and medical abortion were reported as a health issue is in direct contrast to that found in newspapers coverage in the USA, where abortion tends to be covered as a political issue. Woodruff ^53^ analyzed news articles on abortion from select USA newspapers during 2013-2016, a similar time period to our content analysis, and found that abortion was not covered as a health issue but as a political issue, and specifically an electoral issue. The most common abortion content is this American study was the relationship to electoral politics and candidate’s positions on abortion. Their study found only 5.6% of articles identified that abortion restrictions pose barriers to abortion access and 2.8% identified abortion as being safe and common. In contrast, almost half (190, 47.15%) of the articles in our Canadian content analysis identified one or more regulatory, financial and/or health system barriers to medical abortion. Further, 68.2%, n=274 discussed how increased mifepristone access would increase abortion availability, and 11.1% of articles explicitly stated that that medical abortion was safe, with supporting information from the WHO or other reputable scientific bodies.

In contrast to the American findings, we found that only a minority of articles (6.2%) discussed the role of a politician or policy maker’s personal view on abortion, and among these 88% included arguments against personal views influencing public policy. This dramatic difference in the representation of abortion in print media between Canada and the USA may be a reflection of a Canadian political value that public policy should not be driven by personal views. In addition, Farney (2009)^54^ explains that, in contrast to American conservatives, Canadian conservatives have historically distanced themselves from socially conservative party positions on issues such as abortion and lesbian, gay and bisexual rights. Certainly, our content analysis indicates that publicly expressing anti-abortion views is not perceived as an effective electoral strategy by political parties or most elected officials.

We found that the vast majority of Canadian articles included pro-abortion viewpoints, yet abortion as a women’s rights issue was only identified in a minority of articles (15.4%). Media coverage on the introduction and regulation of mifepristone for medical abortion in Canada between 2015 and 2019 reflected the historical demotion of partisanship on social issues in Canada in that the discussion of medical abortion was framed as a health issue, frequently exploring different components necessary to realize the right to health, rather than as moral or political debate which would pit the rights of the fetus against those of the woman.

### Strengths and Limitations

Our content analysis comprehensively included all print media over the time period relevant to mifepristone introduction and regulation, from six months before initial approval until after the last Canadian province included mifepristone in the provincial drug plan. We also included all print media, regardless of circulation number or geographic coverage (national versus local newspaper). This comprehensiveness allowed us to capture the entire narrative presented by the print media and to examine if and how it changed over time as the regulatory landscape evolved. We used an *a priori* coding scheme and identified key themes in advance. The data presented in this paper only considered print news media and did not include radio, television or social media. Our study considered only one facet of media reporting to the public (print media) and therefore cannot be taken as fully representative of the wider media coverage of mifepristone’s introduction and regulation in Canada.

## Conclusion

We found coverage by Canadian print media was supportive of the approval and introduction of mifepristone for medical abortion and presented medical abortion as a health issue framed and supported by science, rather than as an issue of partisan politics, morality or women’s rights. Print media coverage communicated arguments about mifepristone medical abortion’s potential to improve fulfillment of the right to health by advancing availability, accessibility, acceptability and quality. Mifepristone’s introduction in Canada was presented as a way to increase women’s health, specifically within rural areas where disparities in abortion access exist. Speakers with expertise, physicians and academic researchers, were cited frequently in support of increasing the availability, access and acceptability of medical abortion, and their statements and stature reinforced perceptions that mifepristone medical abortion fulfills the quality aspect of the right to health. Anti-medical abortion discourse was marginalized in the overall narrative. The marginalization of anti-abortion arguments is an example of evidentiary balance in reporting, where the weight of expert opinion supports abortion as a safe and necessary health procedure.

This content analysis is not able to draw conclusions about the influence of coverage on political decision-making or the knowledge of the public. However, the Canadian print media presented arguments that were overwhelmingly favorable towards the expansion of mifepristone medical abortion and served to destigmatize abortion by framing the introduction and universal coverage of medical abortion as a health issue and by providing evidence-based information about safety.

## Data Availability

All relevant data are within the manuscript and its Supporting Information files.

## References

1. Dunn S, Cook R. Medical abortion in Canada: behind the times. CMAJ : Canadian Medical Association journal = journal de l’Association medicale canadienne. 2014;186(1):13–14.

2. World Health Organization. 19th WHO Model List of Essential Medicines. Available at: https://www.who.int/topics/essential_medicines/en/ Accessed 2019 Mar 242015.

3. World Health Organization. Abortion care guideline. World Health Organization Guidelines. 2022;Licence: CC BY-NC-SA 3.0 IGO(Available at: https://www.who.int/publications-detail-redirect/9789240039483): Geneva.

4. Grant K. Long-awaited abortion pill Mifegymiso makes Canadian debut. The Globe and Mail2017 Jan 20.

5. Health Canada. Regulatory decision summary: MIFEGYMISO. 2015. Available at https://cart-grac.ubc.ca/np-mifepristone-study/regulatory-decision-summary-sbd_-mifegymiso-2015-health-canada/?login(last Accessed on August 27, 2021). Ottawa, Canada: Government of Canada.

6. Shaw D, Norman WV. When there are no abortion laws: A case study of Canada. Best practice & research Clinical obstetrics & gynaecology. 2020;62:49–62.

7. R. v. Morgentaler. Supreme Court Judgments. 1988-01-28. Case number: 19556., 2019 (Supreme Court Judgments 1988).

8. Canada. Canada Health Act. In: Government of Canada, ed. R.S.C., 1985, c. C-6. Vol c. C-6. Canada 1985.

9. Criminal Law Amendment Act, 1968-69, Chapter 38 1968-69 (Statutes of Canada 1969).

10. Canadian Institute for Health Information. Induced abortions reported in Canada 2018. In: CIHI, ed. Vol 2020. Ottawa, Canada. Available at [https://www.cihi.ca/en/access-data-reports/results?query=abortion&Search+Submit=] Accessed 2020 Jan 25: Government of Canada; 2020.

11. Norman WV. Induced abortion in Canada 1974-2005: trends over the first generation with legal access. Contraception. 2012;85(2):185–191.

12. Sethna C, Doull M. Spatial disparities and travel to freestanding abortion clinics in Canada. Women Stud Int Forum. 2013;38:52–62.

13. Norman WV, Guilbert ER, Okpaleke C, et al. Abortion health services in Canada: Results of a 2012 national survey. Canadian family physician Medecin de famille canadien. 2016;62(4):e209–e217.

14. United Nations. Committee on the Elimination of Discrimination against Women: Concluding observations on the combined eighth and ninth periodic reports of Canada. Available at: http://www.etoconsortium.org/nc/en/404/?tx_drblob_pi1%5BdownloadUid%5D=194, Accessed on June 18, 2021.: United Nations High Commissioner on Human Rights,; 2016.

15. Costescu D, Guilbert E, Bernardin J, et al. Medical Abortion. Journal of obstetrics and gynaecology Canada : JOGC = Journal d’obstetrique et gynecologie du Canada : JOGC. 2016;38(4):366–389.

16. Norman WV, Munro S, Brooks M, et al. Could implementation of mifepristone address Canada’s urban-rural abortion access disparity: a mixed-methods implementation study protocol. BMJ open. 2019;9(4):e028443.

17. Munro S, Guilbert E, Wagner M-S, et al. Perspectives Among Canadian Physicians on Factors Influencing Implementation of Mifepristone Medical Abortion: A National Qualitative Study. Annals of family medicine. 2020;18(5):413–421.

18. Winikoff B, Hassoun D, Bracken H. Introduction and provision of medical abortion: a tale of two countries in which technology is necessary but not sufficient. Contraception. 2011;83(4):322–329.

19. Health Canada. MIFEGYMISO (mifepristone and misoprostol tablets) - Canadian Distribution and Administration Program. In. Available at: http://healthycanadians.gc.ca/recall-alertrappel-avis/hc-sc/2017/63330a-eng.php?_ga=2.209520661.688245650.1504820818-1040716025.1490733726 Accessed 2017 May 19.

20. Health Canada. Mifegymiso. Health Canada updates prescribing and dispensing information for Mifegymiso. 7 Nov 2017. 2017 Nov 7; https://hpr-rps.hres.ca/reg-content/regulatory-decision-summary-detail.php?lang=en&linkID=RDS00294 Accessed 2021 Aug 27.

21. Health Canada. Health Canada approves updates to Mifegymiso prescribing information: Ultrasound no longer mandatory. Recalls and Safety Alerts 2019; https://healthycanadians.gc.ca/recall-alert-rappel-avis/hc-sc/2019/69620a-eng.php. Accessed April 18, 2019.

22. Cook FL, Tyler TR, Goetz EG, et al. Media and Agenda Setting: Effects on the Public, Interest Group Leaders, Policy Makers, and Policy. Public Opin Quart. 1983;47(1):16.

23. Otten AL. The Influence of the Mass Media on Health Policy. Health Affairs. 1992;11(4):111–118.

24. Pan Z, Kosicki G. Framing analysis: An approach to news discourse. Political Communication. 1993;10(1):55–75.

25. World Health Organization. Human rights and health. Available at https://www.who.int/news-room/fact-sheets/detail/human-rights-and-health. x29 December 2017.

26. World Health Organization and Office of the United Nations High Commissioner for Human Rights. The right to health. Fact sheet. Available at https://www.ohchr.org/Documents/Issues/ESCR/Health/RightToHealthWHOFS2.pdf. 2021.

27. Office of the United Nations High Commissioner for Human Rights and World Health Organization. ‘The Right to Health’, Fact Sheet No. 31. Available at https://www.ohchr.org/sites/default/files/Documents/Publications/Factsheet31.pdf. Geneva 2008.

28. ProQuest. Canadian Newsstream. 2021; https://about.proquest.com/en/products-services/canadian_newsstand/. Accessed December 14, 2021.

29. Indigenous Services Canada. Non-insured Health Benefits Program: First Nations and Inuit Health Branch: Annual Report 2017-2018. Nunavut client population [value = 35,079]. Available at [https://www.sac-isc.gc.ca/eng/1581294869253/1581294905909#chp2Statistics]. 2021. Accessed Jul 1, 2022.

30. Statistics Canada. Population estimates, quarterly. Table: 17-10-0009-01 (formerly CANSIM 051-0005). Retrieved from https://doi.org/10.25318/1710000901-eng. 2018. Accessed July 1, 2022.

31. Deachman B. Independents’ Day; Candidates free of burdens of party politics could have a big role to play. The Ottawa Citizen. 2015;2015(October 7):A7.

32. Action Canada for Sexual Health and Rights. FAQ: The Abortion Pill Mifegymiso. Available at https://www.actioncanadashr.org/resources/factsheets-guidelines/2019-04-06-faq-abortion-pill-mifegymiso. (Accessed 2021 Dec 14). 2019; https://www.actioncanadashr.org/resources/factsheets-guidelines/2019-04-06-faq-abortion-pill-mifegymiso. Accessed December 14, 2021.

33. WHO Department of Reproductive Health and Research HRP. Essential Medicines List Application Mifepristone–Misoprostol for Medical Abortion. Geneva: World Health Organization; Oct., Available at: https://www.who.int/selection_medicines/committees/expert/22/applications/s22.1_mifepristone-misoprostol.pdf?ua=1 2018.

34. Kirkey S. Medical body seeks approval of abortion pill in Canada; Drug won’t cause increase in terminated pregnancies, group says. The Vancouver Sun. 2015 Jan 13, 2015.

35. Kirkey S. Approval of Abortion pill sought; Self-administered. National Post. 2015 Jan 13, 2015.

36. Kirkey S. Approval of Abortion pill sought. National Post. 2015 Jan 13, 2015.

37. Kirkey S. Home abortion pill gets doctors’ support. Montreal Gazette. 2015 Jan 13, 2015.

38. Sanders C. Abortion drug boon to rural areas. Winnipeg Free Press. 2015 Jul 31, 2015.

39. Postmedia News. Approve abortion pill mifepristone, doctors group urges Health Canada. National Post2015: News, Canada.

40. Bains C. B.C. latest province to pay for ‘abortion pill’. The Medicine Hat News. 2018 Jan 03, 2018: 6.

41. Clarke CE, Dixon GN, Holton A, McKeever BW. Including “evidentiary balance” in news media coverage of vaccine risk. Health Commun. 2015;30(5):461–472.

42. Purcell C, Hilton S, McDaid L. The stigmatisation of abortion: a qualitative analysis of print media in Great Britain in 2010. Culture, health & sexuality. 2014;16(9):1141–1155.

43. Sisson G, Herold S, Woodruff K. “The stakes are so high”: interviews with progressive journalists reporting on abortion. Contraception. 2017;96(6):395–400.

44. Thomas RJ, Tandoc EC, Hinnant A. False Balance in Public Health Reporting? Michele Bachmann, the HPV Vaccine, and “Mental Retardation”. Health Communication. 2017;32(2):152–160.

45. Raymond EG, Grimes DA. The comparative safety of legal induced abortion and childbirth in the United States. Obstetrics and gynecology. 2012;119(2 Pt 1):215–219.

46. World Health Organization. Global Abortion Policies Database. 2018; https://abortion-policies.srhr.org/.

47. Sethna C, Doull M. Far From Home? A pilot study tracking women’s journeys to a Canadian abortion clinic. 2007;29(8):640–647.

48. Sethna C, Palmer B, Ackerman K, Janovicek N. Choice, Interrupted: Travel and Inequality of Access to Abortion Services since the 1960s. Labour-Travail. 2013;71(71):29-+.

49. Ferris LE, McMain-Klein M, Iron K. Factors influencing the delivery of abortion services in Ontario: a descriptive study. Family planning perspectives. 1998;30(3):134–138.

50. Kaposy C. Improving abortion access in Canada. Health care analysis : HCA : journal of health philosophy and policy. 2010;18(1):17–34.

51. Nelson E. Autonomy, Equality, and Access to Sexual and Reproductive Health Care Special Issue: Health Law. Alberta Law Review. 2016;54:707–726.

52. Norman WV, Hestrin B, Dueck R. Access to Complex Abortion Care Service and Planning Improved through a Toll-Free Telephone Resource Line. Obstet Gynecol Int. 2014;2014:913241.

53. Woodruff K. Coverage of Abortion in Select U.S. Newspapers. Women’s Health Issues. 2019;29(1):80–86.

54. Farney J. The Personal Is Not Political: The Progressive Conservative Response to Social Issues. American Review of Canadian Studies. 2009;39(3):242–252.

